# Predictors of postoperative biochemical remission in acromegaly

**DOI:** 10.1101/2020.06.26.20141325

**Authors:** Shun Yao, Wen-Li Chen, Sherwin Tavakol, Farhana Akter, Michael P. Catalino, Jie luo, Xiao-Peng Guo, Ai-Liang Zeng, Leo Zekelman, Zhi-Gang Mao, Yong-Hong Zhu, Qing-Zhi Wu, Edward R Laws, Wenya Linda Bi, Hai-Jun Wang

## Abstract

**OBJECTIVE:** Acromegaly is a rare neuroendocrine condition that can lead to significant morbidity; therefore, large studies are invaluable for understanding the disease burden. Despite China’s vast population size, studies on acromegaly remain sparse. In this report, we aimed to investigate clinical characteristics and predictors of biochemical remission after surgery for acromegaly.

**METHODS:** A retrospective nationwide study was conducted using patient-reported data from the China Acromegaly Patient Association (CAPA) from 1998 to 2018. Univariate analyses were performed using Wilcoxon rank-sum tests, two-sample t-tests, and chi-squared tests. Using the purposeful selection method, multivariate logistic regression analysis was employed to determine independent predictors of biochemical remission at 3 months in patients after surgery.

**RESULTS:** Of the 575 cases (mean age: 37.3 years; 59% female), macroadenomas and invasive tumors (Knosp score 3-4) were 87% and 61%, respectively. Ninety-five percent of patients were treated first with surgery (5.1% transcranial and 94.9% endonasal) and 38.3% exhibited biochemical remission at 3-months postoperatively. The following independent predictors of biochemical remission were identified: preoperative growth hormone (GH) levels between 12 and 28 μg/L [odds ratio (OR)=0.61; 95% confidence interval (CI), 0.39-0.96; p=0.031], preoperative GH levels >28 μg/L (OR=0.56; 95% CI, 0.35-0.90; p=0.016), macroadenoma (OR=0.57; 95% CI, 0.33-0.97; p=0.041), giant adenomas (OR=0.17; 95% CI, 0.06-0.44; p=0.0005), Knosp score 3-4 (OR=0.39; 95% CI, 0.25-0.59; p<0.0001), and preoperative medication usage (OR=2.16; 95% CI, 1.38-3.39; p=0.0008).

**CONCLUSIONS:** In this nationwide study spanning over two decades, we highlight that higher preoperative GH levels, large tumor size, and greater extent of tumor invasiveness are associated with a lower likelihood of biochemical remission at 3-months after surgery, while preoperative medical therapy increases the chance of remission.

## Introduction

Acromegaly is a rare condition characterized by the overproduction of growth hormone (GH) from a hypersecreting pituitary tumor.^1–3^ Clinical manifestations of GH-secreting tumors can manifest insidiously and result in a delayed diagnosis, while the systemic repercussions of insulin-like growth factor 1 (IGF-1) excess, resultant from GH stimulation, are associated with increased morbidity and mortality.^4^ Symptoms and signs of acromegaly include facial changes, excessive skeletal growth, soft tissue hypertrophy, obstructive sleep apnea, hyperhidrosis, headache, and visual impairments.^2,5–7^ Persistent high levels of GH and/or IGF-1 may also result in glucose intolerance, osteoporosis, arthritis, reproductive disorders, and cardiovascular dysfunction, ultimately driving premature mortality.^2,4–7^

Given the rarity of acromegaly, large studies that characterize the disease pattern, describe risk factors predictive of poor disease control and inform management decisions of this condition are scarce. In particular, there is a notable gap in data regarding prevalence, disease severity, treatments administered, and clinical outcomes for acromegaly patients in China. We present the first report from the China Acromegaly Patient Association (CAPA), which is a non-profit, patient-advocacy organization with a patient-reported database encompassing treatment from 112 hospitals. Using data from this directory, we aim to provide a comprehensive description of clinical characteristics, and to identify predictors of biochemical remission at 3 months after surgery in patients with acromegaly in China, so as to improve recognition for the diagnosis of acromegaly and to guide management decisions.

## Methods

### Study Design and Data Collection

We retrospectively reviewed the records of 916 acromegaly patients enrolled into the multi-institutional China Acromegaly Patient Association (CAPA) database (https://capa.wohenok.com) between June 1998 and December 2018. The CAPA database is patient-reported and relies on the patient submission of demographic and clinical information following the diagnosis and treatment of acromegaly, drawing upon patient information from 112 hospitals across China (**Supplemental Materials S1**). Patients lacking data on treatment interventions or biochemical status at follow-up were excluded from the analysis, resulting in a final cohort of 575 patients.

We extracted demographic data (including age, sex, location of residence, and insurance type), disease duration, clinical symptoms, biochemical status (at diagnosis and on follow-up), radiographic features, treatment history, histopathology, postoperative complications, and follow-up at 3 months after treatment. Approval (approved ID [2020] 091) for this retrospective study was obtained from CAPA and the institutional review board of The First Affiliated Hospital of Sun Yat-sen University, Guangzhou, China.

### Variables Analyzed

All patients in the CAPA database were diagnosed with acromegaly in accordance with established guidelines (**Box 1**).^8^ Diagnostic factors include classic signs and symptoms, biochemical assays, and radiographic identification of a pituitary tumor.

#### **Box 1** | **Diagnostic and biochemical remission criteria for acromegaly**

**Diagnostic criteria**

- Classic signs and symptoms of acromegaly
- Endocrinological assay: fasting growth hormone (GH) > 2.5 μg/L, glucose GH inhibition test with lowest GH > 1 μg/L, and insulin-like growth factor 1 (IGF-1) level higher than the normal range of age- and sex-matched healthy individuals
- Radiographic identification of a pituitary tumor on computed tomography (CT), high-resolution magnetic resonance imaging (MRI), or dynamic contrast-enhanced MRI of the sellar region
- Pituitary hormone aberrancies: abnormal levels in prolactin (PRL), follicle stimulating hormone (FSH), luteinizing hormone (LH), thyroid-stimulating hormone (TSH), and/or adrenocorticotropic hormone (ACTH)

**Biochemical remission criteria ^8^**

- Random serum GH < 2.5 μg/L, glucose GH inhibition test with GH < 1 μg/L
- Normalized serum IGF-1 level

### Radiographic features

Imaging was reviewed by either an attending neurosurgeon or neuroradiologist. Cavernous sinus invasion was classified using the Knosp Grading System.^9^ Tumor invasion was defined as Knosp grades 3 & 4 (evidence of tumor crossing the lateral border of the cavernous carotid artery on coronal MRI image), while lack of invasion was defined as a Knosp score of 0 to 2 (margin of tumor medial to the lateral border of cavernous carotid artery).^9,10^

Tumor size was classified into three categories: microadenomas (maximum diameter <1 cm), macroadenomas (1-4 cm), and giant adenomas (≥4 cm).^11^

### Location of residence

The geographic grouping was performed based on the Yicai Media Group (https://www.yicai.com) stratification of Chinese cities and towns into 5 tiers, which we adapted to classify patient residence into the following categories based on population density and size of economy: large city, medium city, and small city/town.

### Insurance coverage

There are three insurance schemes in China, including the urban employee-based basic medical insurance scheme (UEBMI; launched in 1998), urban resident-based basic medical insurance scheme (URBMI; launched in 2007), and rural new cooperative medical scheme (NCMS; launched in 2003), which is intended to provide affordable health insurance for the poor and improve the equity of health services in rural China.^12^ In general, UEBMI covers a broader array of medical services than URBMI, with NCMS having the least robust coverage.

### Treatment

Primary treatment modalities included medication, radiotherapy, or surgery, which comprised of microscopic endonasal, endoscopic endonasal, or transcranial surgery (**Figure 1**). The receipt of preoperative medication was decided by an agreement between the physicians and the patients, according to the established guidelines for acromegaly management^3,8^ and the coverage of their medical insurance or financial capacity. All surgical specimens were analyzed using immunohistopathology. Patients who did not achieve initial biochemical remission received one of the following secondary treatments: repeat surgery, radiotherapy, medication, or combined-therapy.

**Figure 1.**
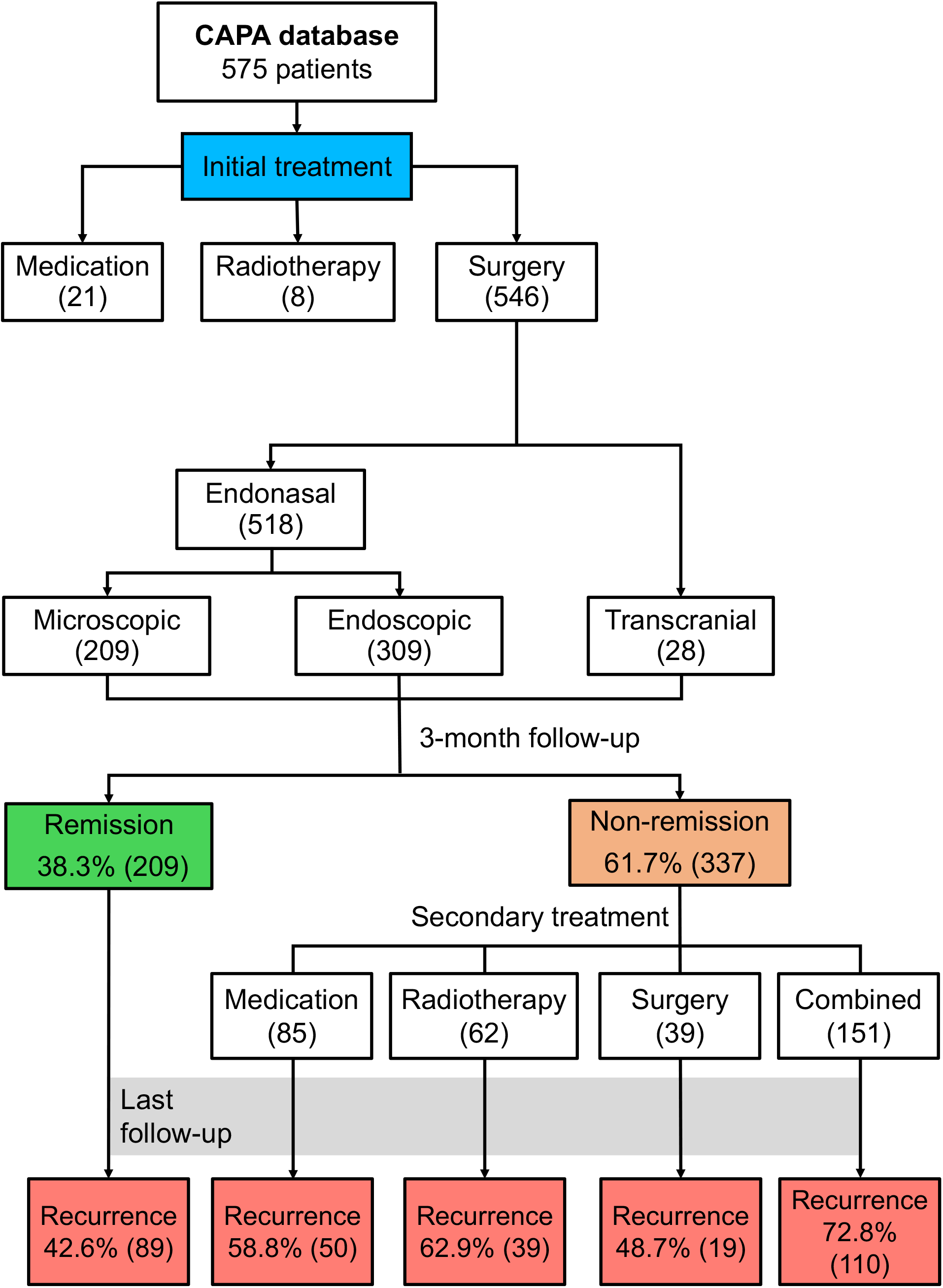
Treatment pathways among patients in the CAPA cohort.

### Outcome Measures

The primary outcome of interest was biochemical remission at 3 months postoperatively. The recurrence rate was also identified using the latest follow up information. The criteria for postoperative biochemical remission after surgery for acromegaly were based on the established guidelines (**Box 1**).^8^

### Statistical Analysis

Baseline characteristics were described using mean ± standard deviation (SD), median (interquartile range, IQR), or frequency according to the variable type. Group differences were compared using the two-sample Student’s t-test or Mann-Whitney U-test for continuous variables, and Pearson’s chi-squared test (two-tailed) or Fisher’s exact test for categorical variables, as appropriate. The family discovery rate (FDR) correction was performed across all univariate analyses to adjust for multiple testing. A purposeful selection approach to multivariate logistic regression analysis was performed to determine the independent predictors of biochemical remission at 3 months postoperatively in patients with acromegaly. Results were presented as odds ratios (ORs) with 95% confidence intervals (CI). The collinearity between variables was explored and evaluated by the variance influence factor (VIF) in the regression analysis. Significance was set at *p* < 0.05 (two-tailed). All statistical analyses were performed using statistical packages in R (https://www.R-project.org/).^13^

## Results

### Cohort

We retrospectively analyzed presenting features, imaging, and clinical outcomes for 575 acromegaly patients (mean age 37.3±10.3 years; 59% women) from the multi-institutional patient-reported CAPA database. The median time from symptom onset to diagnosis was 5 years (range: 1-10 years). The most common clinical symptoms and signs at presentation were enlarged nose and prognathism (97.4%) (**Table 1**). Visual field abnormalities, resulting from tumor mass effect, occurred in 49.2% of patients. Apoplexy occurred in 9.6%, producing tumor mass effect. Other comorbidities associated with acromegaly were reported as follows: cardiovascular disease (40.0%), impaired glucose tolerance (31.0%), osteoporosis (22.6%), colonic polyps (16.3%), arthritis (15.8%), and colon cancer (0.7%).

**Table 1.** Demographics and clinical presentation (N=575)

| Categories | Distribution |
| --- | --- |
| **Mean age** (years), mean ± SD | 37.3 ± 10.3 |
| **Male / Female** | 235/340 |
| **Disease duration** (years), median (IQR) | 5 (2.5-8.0) |
| **Follow-up after initial diagnosis** (months), median (IQR) | 54 (30-86) |
| **Follow-up after initial treatment** (months), median (IQR) | 32 (13-61) |
| **Clinical symptoms and signs, n (%)** |  |
| *Enlarged nose and prognathism* | 560 (97.4) |
| *Overgrowth of extremities* | 537 (93.4) |
| *Menstrual disturbance (female)* | 227 (66.8) |
| *Snoring or sleep apnea* | 358 (62.3) |
| *Headache* | 336 (58.4) |
| *Hyperhidrosis* | 323 (56.2) |
| *Hypertrophy of frontal bones* | 287 (49.9) |
| *Visual field abnormalities* | 283 (49.2) |
| *Cardiovascular disease* | 230 (40.0) |
| *Impaired glucose tolerance* | 178 (31.0) |
| *Hypertrichosis* | 197 (34.3) |
| *Jaw malocclusion* | 139 (24.2) |
| *Osteoporosis* | 130 (22.6) |
| *Colonic polyps* | 94 (16.3) |
| *Arthritis* | 91 (15.8) |
| *Galactorrhea* | 88 (15.3) |
| *Colonic cancer* | 4 (0.7%) |
| **Resident area** |  |
| *Large city* | 252 (43.8) |
| *Medium city* | 206 (35.8) |
| *Small city/town* | 117 (20.4) |
| **Insurance type** |  |
| *Urban Employee Basic Medical Insurance* | 366 (63.7) |
| *New Cooperative Medical Scheme* | 130 (22.6) |
| *Urban Resident Basic Medical Insurance* | 79 (13.7) |

**Abbreviations:** IQR, interquartile range; SD, standard deviation.

Large and giant tumors dominated this cohort, with 73.2% being macroadenomas, 10.1% being giant adenomas, and 16.7% being microadenomas (**Table 2, Figure 2A**). Consistent with the prevalence of large tumors in this cohort, two-thirds of the tumors demonstrated cavernous sinus invasion (**Table 2, Figure 2A**).

**Table 2.**
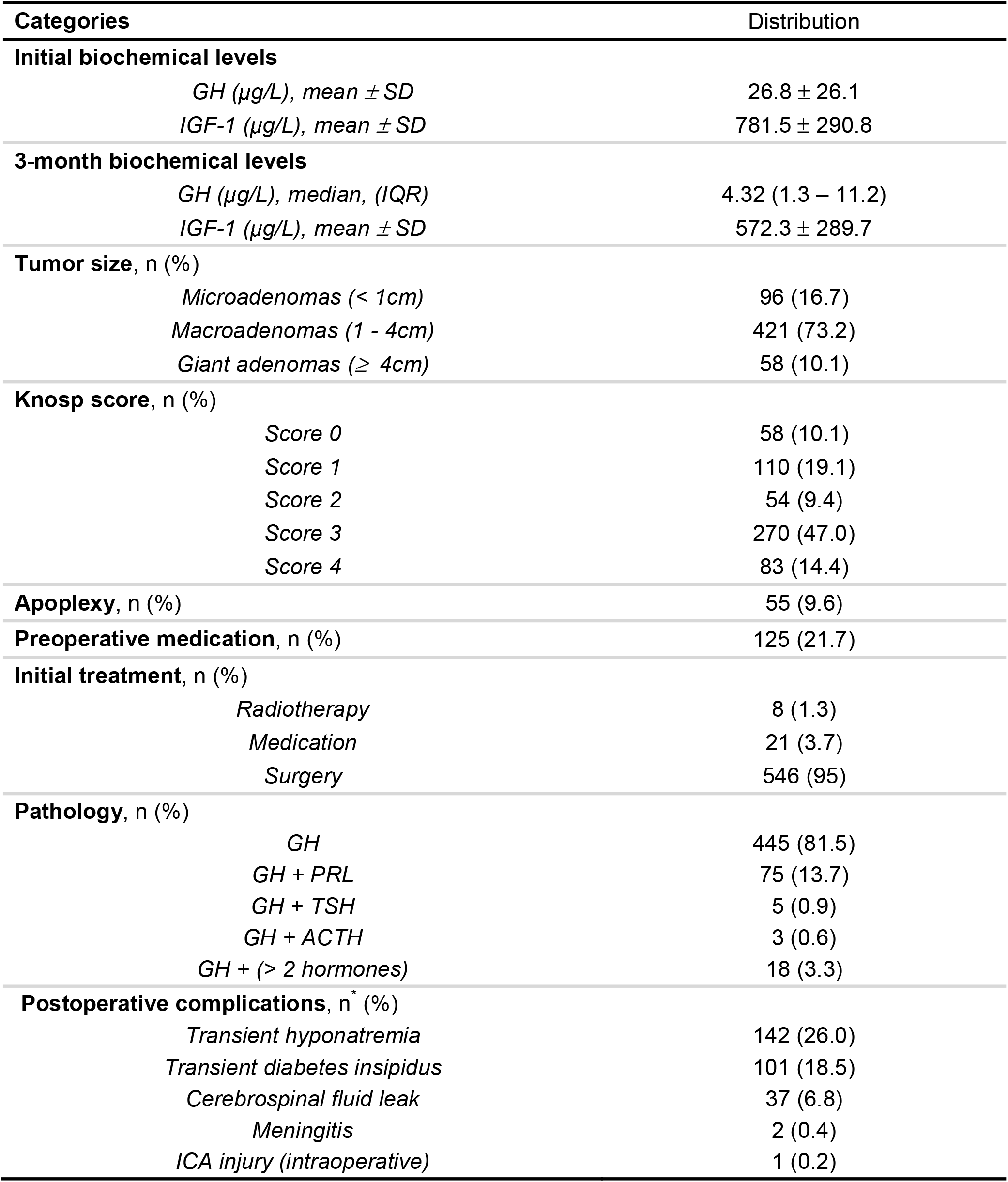
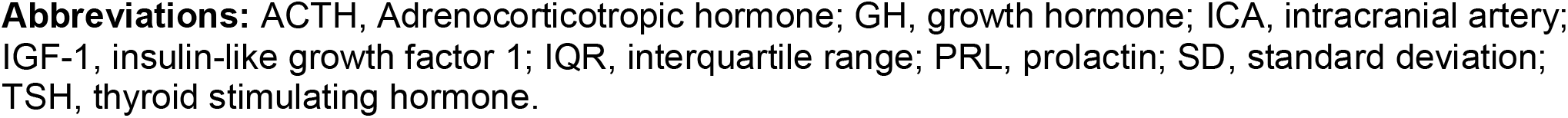
Tumor characteristics and treatment (N=575)

| Categories | Distribution |
| --- | --- |
| **Initial biochemical levels** |  |
| *GH (μg/L), mean ± SD* | 26.8 ± 26.1 |
| *IGF-1 (μg/L), mean ± SD* | 781.5 ± 290.8 |
| **3-month biochemical levels** |  |
| *GH (μg/L), median, (IQR)* | 4.32 (1.3 – 11.2) |
| *IGF-1 (μg/L), mean ± SD* | 572.3 ± 289.7 |
| **Tumor size, n (%)** |  |
| *Microadenomas (< 1cm)* | 96 (16.7) |
| *Macroadenomas (1 - 4cm)* | 421 (73.2) |
| *Giant adenomas (≥ 4cm)* | 58 (10.1) |
| **Knosp score, n (%)** |  |
| *Score 0* | 58 (10.1) |
| *Score 1* | 110 (19.1) |
| *Score 2* | 54 (9.4) |
| *Score 3* | 270 (47.0) |
| *Score 4* | 83 (14.4) |
| **Apoplexy, n (%)** | 55 (9.6) |
| **Preoperative medication, n (%)** | 125 (21.7) |
| **Initial treatment, n (%)** |  |
| *Radiotherapy* | 8 (1.3) |
| *Medication* | 21 (3.7) |
| *Surgery* | 546 (95) |
| **Pathology, n (%)** |  |
| *GH* | 445 (81.5) |
| *GH + PRL* | 75 (13.7) |
| *GH + TSH* | 5 (0.9) |
| *GH + ACTH* | 3 (0.6) |
| *GH + (> 2 hormones)* | 18 (3.3) |
| **Postoperative complications, n\* (%)** |  |
| *Transient hyponatremia* | 142 (26.0) |
| *Transient diabetes insipidus* | 101 (18.5) |
| *Cerebrospinal fluid leak* | 37 (6.8) |
| *Meningitis* | 2 (0.4) |
| *ICA injury (intraoperative)* | 1 (0.2) |

**Abbreviations:** ACTH, Adrenocorticotrophic hormone; GH, growth hormone; ICA, intracranial artery; IGF-1, insulin-like growth factor 1; IQR, interquartile range; PRL, prolactin; SD, standard deviation; TSH, thyroid stimulating hormone.

**Figure 2.**
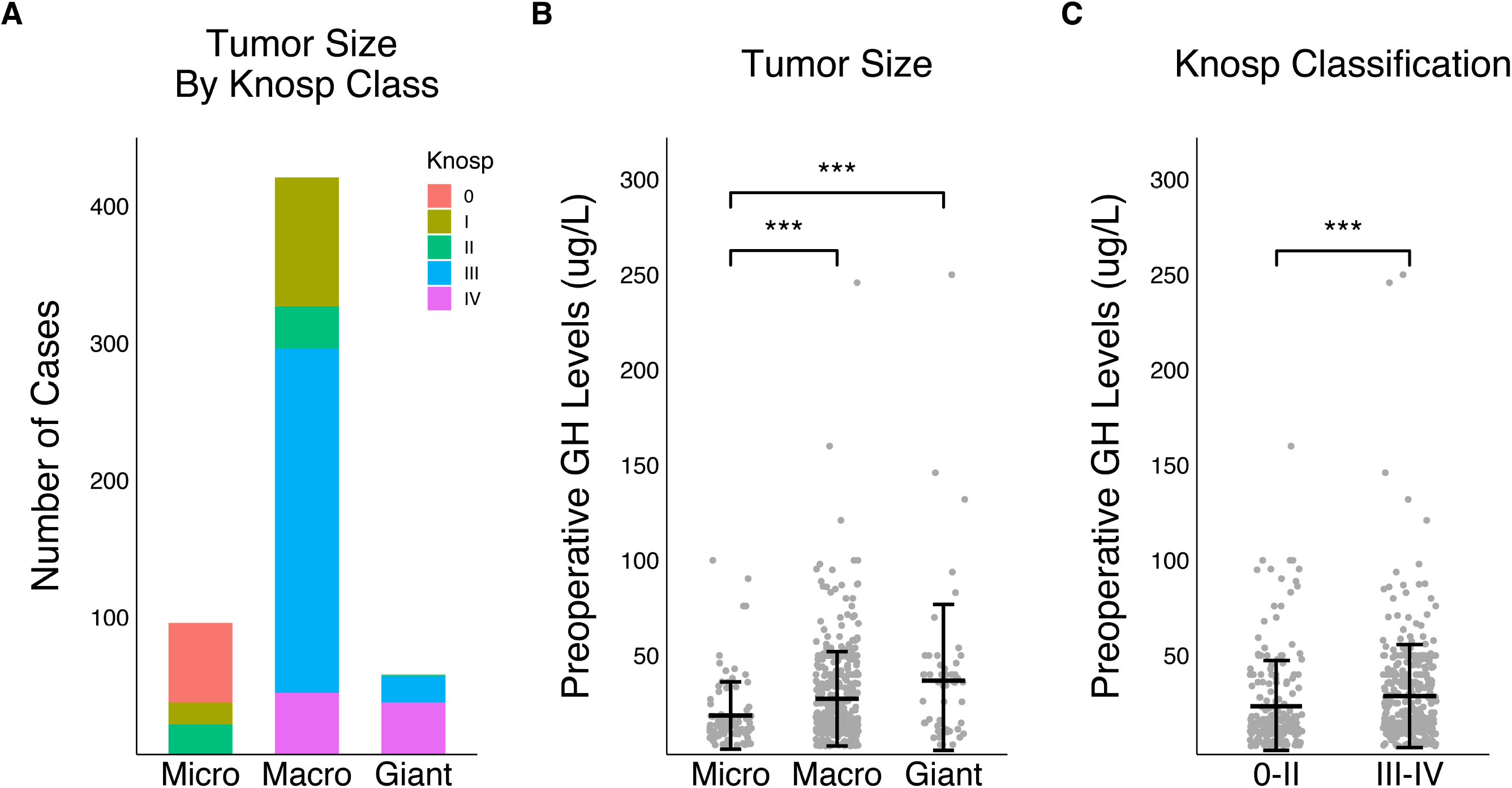
(A) Distribution of cases by Knosp classification and tumor size. (B & C) The association of preoperative growth hormone (GH) levels compared with tumor size and Knosp score.

The majority of patients lived in large cities (43.8%), while only 20.4% of acromegaly patients were from small cities/towns. The urban employer-driven UEBMI scheme was the most prevalent insurance coverage in our cohort (63.7%), followed by the urban resident NCMS (22.6%) and the rural URBMI (13.7%) plans (**Table 1**).

Median follow-up after the initial diagnosis was 54 months (range 6-295 months), while median follow-up after initial treatment was 32 months (range 3-256 months), suggesting an average of 19 months interval delay between diagnosis and treatment.

### Endocrinological Status

Among 575 acromegaly patients, the mean levels of GH and IGF-1 prior to surgery were 26.8 μg/L and 781.5 μg/L respectively. Mean preoperative GH level was significantly higher in patients with macroadenomas (27.3 μg/L) and giant adenomas (36.8 μg/L) compared to those with microadenomas (18.5 μg/L) (both p<0.001) (**Figure 2B**). Additionally, tumors with cavernous sinus invasion exhibited significantly higher median GH levels than those without invasion (28.7 μg/L vs. 23.3 μg/L, p<0.001) (**Figure 2C**). Tumors with plurihormonal expression showed higher median GH levels (22.7 μg/L) than dual-staining pituitary tumors of GH and prolactin (DSPTs) (20.0 μg/L) and GH alone (16.5 μg/L), but these differences did not reach a statistical significance.

### Treatment

546 patients (95%) received surgery as their initial treatment, while 21 patients (3.7%) received medical therapy alone and 8 patients (1.3%) received upfront radiation (**Figure 1**). The most common operative approach was endoscopic endonasal transsphenoidal (94.9%), with transcranial surgery (5.1% of cases) mostly reserved for larger tumors with a greater extent of invasiveness. The mean age of patients who received either medication or radiation only was 49 years (range 32-70 years), compared to a mean of 37 years (range 16-76 years) in patients who underwent surgery (p<0.001). For patients receiving medication alone, mean GH levels trended lower than that of patients who underwent surgery (19.7 vs. 27.0 μg/L, respectively; p=0.036).

### Surgical Outcome

Among the 546 patients who underwent initial surgical resection, histopathology confirmed immunopositivity for GH alone in 81.5%, concurrent GH and prolactin staining in 13.7%, and the presence of additional hormone expression along with GH in the remaining (4.8%) (**Table 2**). Tumors that stained for both GH and prolactin were more likely than solely GH-staining tumors to be invasive (78.7% vs. 61.1%, respectively; p=0.003).^14^ Moreover, the most common postoperative complication was postoperative hyponatremia (sodium <135 mmol/L) (26.0%), followed by diabetes insipidus (18.5%), cerebrospinal fluid leak (6.8%), meningitis (0.4%). One case of internal carotid artery (ICA) injury was reported.

At three months after surgery, 209 (38.3%) patients had achieved biochemical remission (**Table 3, Figure 1**). More aggressive tumors were associated with a lower likelihood of remission at 3 months. Specifically, higher pre-operative GH levels, larger tumors, and the presence of cavernous sinus invasion were all significantly associated with lack of remission (p=0.003, p<0.001, p<0.001, respectively). Although biochemical remission was observed in 63.6% of microadenoma patients, it was only observed in 12.3% of patients with giant adenomas. Similarly, 57.1% of cases with Knosp grade 0-2 experienced remission, compared to only 27.7% of Knosp grade 3-4 tumors.

**Table 3.** Comparison of demographics and clinical presentation between 3-month non-remission and remission cases.

| Categories | Surgical remission |  | P-value <sup>d</sup> |
| --- | --- | --- | --- |
|  | No (n = 337) | Yes (n = 209) |  |
| **Age** (years), mean ± SD | 36.0 ± 9.7 | 38.2 ± 10.1 | 0.08 |
| **Sex** |  |  | 0.966 |
| *Male* | 137 (62.0) | 84 (38.0) |  |
| *Female* | 200 (61.5) | 125 (39.5) |  |
| **Disease duration** (years), median (IQR) | 4 (2 - 8) | 5 (2-8) | 0.361 |
| **Initial GH levels** (μg/L), median (IQR) | 20.0 (12.0 – 40.0) | 14.6 (10.2 – 29.9) | **0.003** |
| **Tumor size**, n (%) |  |  | **< 0.001** |
| *Microadenomas* | 32 (36.4) | 56 (63.6) |  |
| *Macroadenomas* | 255 (63.6) | 146 (36.4) |  |
| *Giant adenomas* | 50 (87.7) | 7 (12.3) |  |
| **Knosp score**, n (%) |  |  | **< 0.001** |
| 0-2 | 84 (42.9) | 112 (57.1) |  |
| 3-4 | 253 (72.3) | 97 (27.7) |  |
| **Apoplexy**, n (%) | 31 (62.0) | 19 (38.0) | 0.966 |
| **Preoperative medication**, n (%) | 67 (53.6) | 58 (46.4) | 0.08 |
| **Surgical approach**, n (%) |  |  | 0.262 |
| *Transcranial surgery* | 22 (78.6) | 6 (21.4) |  |
| *Microscopic TSS* | 131 (62.7) | 78 (37.3) |  |
| *Endoscopic TSS* | 184 (59.5) | 125 (40.5) |  |
| **Pathology**, n (%) |  |  | 0.727 |
| *GH* | 269 (60.5) | 176 (39.5) |  |
| *GH + ACTH* | 2 (66.7) | 1 (33.3) |  |
| *GH + PRL* | 49 (65.3) | 26 (34.7) |  |
| *GH + TSH* | 3 (60.0) | 2 (40.0) |  |
| *GH + (> 2 hormones)* | 14 (77.8) | 4 (22.2) |  |
| **City type of residence** |  |  | 0.521 |
| *Large city* | 138 (58.7) | 97 (41.3) |  |
| *Medium city* | 131 (65.5) | 69 (34.5) |  |
| *Small city/town* | 68 (61.3) | 43 (38.7) |  |
| **Insurance type** |  |  | 0.721 |
| *Urban Employee Basic Medical Insurance* | 211 (61.9) | 130 (38.1) |  |
| *New Cooperative Medical Scheme* | 83 (64.3) | 46 (35.7) |  |
| *Urban Resident Basic Medical Insurance* | 43 (56.6) | 33 (43.4) |  |

<sup>a</sup> **FDR correction**
**Abbreviations:** ACTH, Adrenocorticotrophic hormone; eTSS, endonasal transsphenoidal surgery; GH, growth hormone; IQR, interquartile range; PRL, prolactin; SD, standard deviation; TSH, thyroid stimulating hormone.

**Table 4.** Multivariate predictors of 3-month surgical remission in acromegaly patients.

| Variable | Logistic regression analysis |  |  |
| --- | --- | --- | --- |
|  | OR | 95% CI | P value |
| **Preoperative GH Level (µg/L)<sup>a</sup>** |  |  |  |
| 12 – 28 | 0.61 | 0.39 - 0.96 | **0.031** |
| > 28 | 0.56 | 0.35 - 0.90 | **0.016** |
| **Tumor size<sup>b</sup>** |  |  |  |
| *Macroadenoma* | 0.57 | 0.33 - 0.97 | **0.041** |
| *Giant adenoma* | 0.17 | 0.06 - 0.44 | **0.0005** |
| **Knosp score: 3-4<sup>c</sup>** | 0.39 | 0.25 - 0.59 | **< 0.0001** |
| **Preoperative medication usage** | 2.16 | 1.38 - 3.39 | **0.0008** |

<sup>a</sup>Reference level <12 µg/L
<sup>b</sup>Reference: Microadenoma
<sup>c</sup>Reference: Knosp score 0-2
**Abbreviations:** CI, confidence interval; GH, growth hormone; OR, odds ratio.

Multivariate logistic regression analysis showed that preoperative GH levels >28 μg/L (highest tertile of our cohort) compared to pre-operative GH level <12 μg/L (lowest tertile in our cohort) decreased the odds of remission at 3 months after surgery by 44% (p=0.016), adjusted for tumor size and receipt of preoperative medication. Additionally, having a macroadenoma or giant adenoma decreased the odds of remission by 43% (p=0.041) and 83% (p=0.0005), respectively, compared to having a microadenoma. Similarly, tumors with a Knosp score of 3-4 had 61% less odds of biochemical remission at 3 months than those with a Knosp score of 0-2 (p<0.0001). Adjusted for age, preoperative GH level, tumor size, and Knosp score, preoperative medication usage more than doubled the odds of biochemical remission (OR=2.16, p=0.0008) (**Figure 3**). The reduction of statistical power through a high correlation between two or more explanatory variables (multicollinearity) was not observed in our regression model (maximum VIF=1.3).

**Figure 3.**
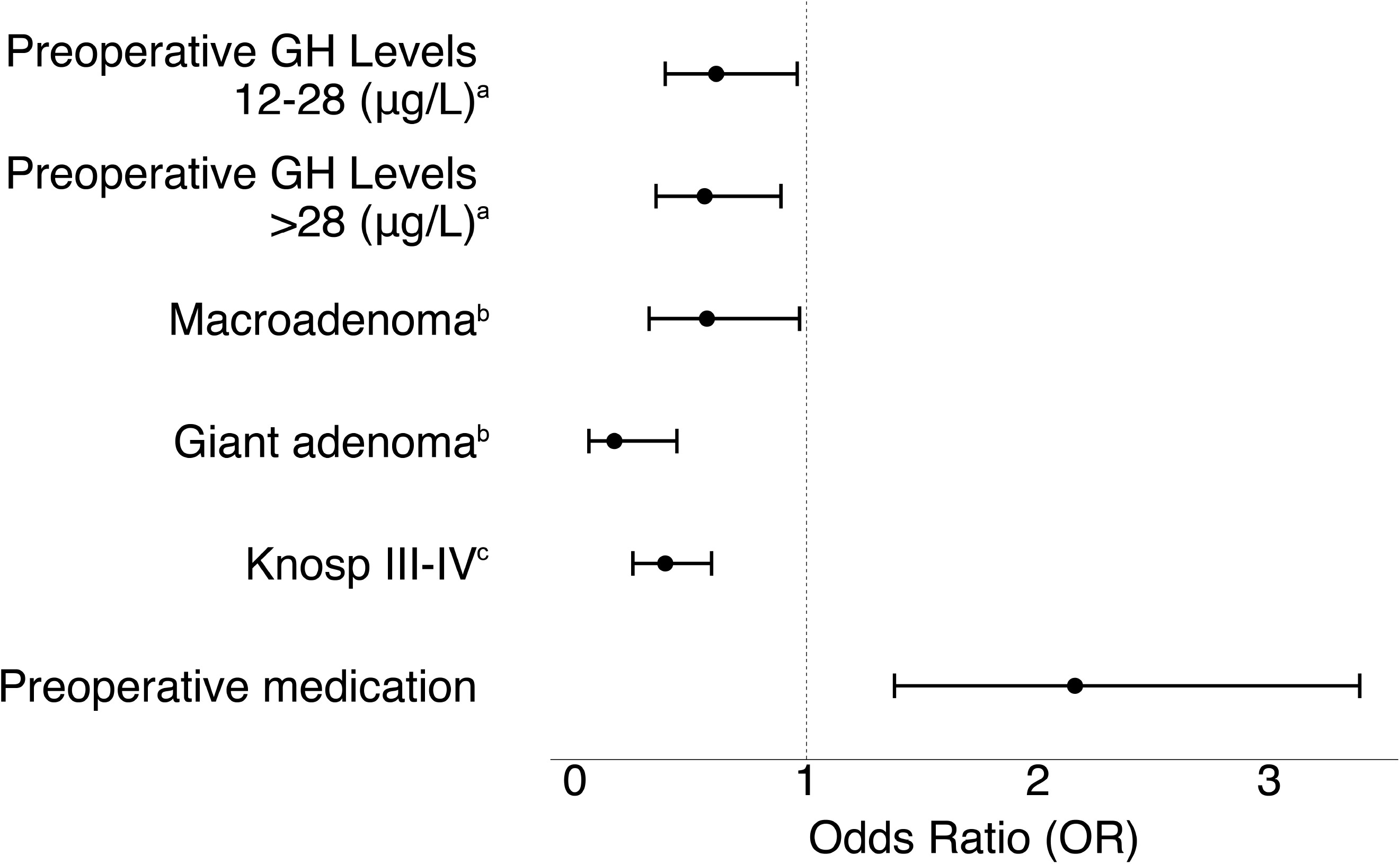
Adjusted odds ratios (OR) and 95% confidence intervals for predictors of biochemical remission at 3 months (p<0.05). ^a^Reference level: <12 μg/L; ^b^Reference: Microadenoma; ^c^Reference: Knosp score 0-2.

The recurrence rate was lower in patients with initial biochemical remission (45.0%) compared to patients without initial remission (65.9%) who presented lowest recurrent rate in repeat surgery (48.6%) compared to patients undergoing radiotherapy (59.6%), medication (63.2%), and combined treatments (75.6%) (**Figure** 1).

Patient age, sex, disease duration, city type of residence, insurance type, apoplexy, surgical approach (microscopic endonasal, endoscopic endonasal, or transcranial), and plurihormonal expression were not significantly associated with postoperative biochemical remission in this cohort.

## Discussion

We present the clinical characteristics of over 500 acromegaly patients in China and observe tumor and treatment factors strongly associated with disease control at three months after surgery or other definitive treatment. Our study represents the largest analysis of clinical features and postoperative outcomes of acromegaly in China, and also constitutes the first application of the CAPA database for clinical research. We observed a 3-month postoperative remission rate of 38.3%, with the likelihood of disease control strongly influenced by the preoperative GH level, tumor size, and tumor invasiveness as indicated by the Knosp grade.

Despite China’s vast population size, medical studies on acromegaly remain sparse, particularly in English-language journals. The earliest record of a surgical case of acromegaly in China dates back to 1929, when the first patient was operated on using transcranial surgery at the Peking Union Medical College Hospital (records endure in archives of the hospital library). Early published studies from China were mostly case series and smaller cohorts. More recently, a single-institution retrospective analysis of 358 Chinese acromegalic patients found the GH remission rate to be 37.3% in patients with invasive macroadenomas without receipt of preoperative somatostatin analogs as compared to 56.4% among those with pre-operative medical treatment.^15^ We drew upon the power of a patient-reported database to expand on the clinical experience in the diagnosis and treatment of acromegaly in China across multiple institutions.

### Clinical characteristics

Although the most commonly reported physical changes in our cohort were similar to previously published series,^2,16^ the detection of such changes may occur in a delayed fashion given the typically insidious onset over decades. Additionally, dental malocclusion, median thenar neuropathy (carpal tunnel syndrome), change in shoe size or ring size, and skin tags may be overlooked signs of acromegaly. Sleep apnea, headache, and hyperhidrosis were observed in over half of patients, in addition to menstrual disturbances in two-thirds of female patients. These, among other common non-physical, symptoms should prompt closer inspection for physical signs of acromegaly and consideration for endocrinological evaluation, including assaying IGF-1 determinations.

Although visual field deficit clearly triggers an alarm for a compressive etiology involving the optic apparatus, 52.5% of patients with macroadenomas did not report any subjective visual impairment. Clinical concern for a large pituitary tumor, as in aggressive acromegaly cases, should prompt formal neuro-ophthalmologic evaluation.

Acromegaly is associated with premature mortality.^17,18^ Older age at diagnosis, increased time from symptom onset to diagnosis, higher GH levels after treatment, and cardiovascular risk factors are independent predictors of early mortality.^18^ Forty percent of patients in this cohort reported cardiovascular symptoms or disease, including heart palpitations, arrhythmias (e.g., atrial fibrillation and heart block), structural heart diseases (e.g., cardiomyopathy and valvular disease), and hypertension. These findings, when viewed in light of the fact that the mean age of our cohort was only 37.3 years, underscores the strikingly significant impact of acromegaly on cardiovascular health.^19^ Furthermore, the high incidence of impaired glucose tolerance (diabetes mellitus), osteoporosis, colonic polyps, and arthritis/arthropathies observed also supports the need for diligent and careful screening by providers, as outlined by the Endocrine Society Clinical Practice Guidelines for acromegaly.^1^ Early diagnosis in conjunction with effective treatment and persistent control/screening of comorbidities are paramount to decreasing the morbidity and mortality associated with this condition.

### Factors influencing biochemical remission

The 38.3% biochemical remission rate at 3-months after surgery for a GH secreting tumor in our cohort is relatively low compared to other reports in the literature.^20,21^ This may reflect the high proportion of large and invasive tumors encompassed by our cohort. Additionally, cultural factors and health literacy for neuroendocrinological diseases may contribute to a delay in seeking medical care, resulting in the disease progression. In addition, 1 in 5 patients in our cohort was from a small city or town, which may further delay access to medical care and endocrinological evaluation. Although we did not observe a statistically significant impact of socioeconomic or insurance status on the likelihood of biochemical remission after surgery for acromegaly in this patient-reported cohort, further investigations are merited to improve the delay between diagnosis and treatment, and also potential delays in the initial recognition of acromegaly, especially amongst individuals with geographical and financial barriers.

Preoperative clinical and biochemical characteristics could impact the likelihood of biochemical remission, including age at diagnosis,^22^ gender,^22^ preoperative serum levels of GH and IGF-1,^20,23–27^ tumor size,^20,24,28,29^ preoperative medication usage,^15,21^ tumor invasiveness,^20,24,27–31^ and histological pathology.^24,30^ Similar to other studies, our analysis also revealed that larger tumors (giant and macroadenomas) were less likely than small tumors to enter biochemical remission at 3 months. Further, patients with invasive tumors and higher preoperative GH levels were less likely to attain remission, adjusted for tumor size and receipt of preoperative medical therapy. Given that aggressive tumors were shown to be less likely to enter biochemical remission postoperatively, early surgical intervention should be instituted to halt further progression of disease.^32^

Pharmaceutical treatment, including somatostatin receptor ligands (SRLs), growth hormone receptor antagonists, and dopamine agonists (DA), have shown to have a beneficial impact on surgical outcomes as both neoadjuvant and adjuvant therapy,^15,21^ which was also demonstrated by our team.^33^ In particular, administration of somatostatin analogs prior to surgery is associated with higher endocrinological remission than adjuvant treatment alone.^15,34–37^ Likewise, the use of preoperative medical treatment more than doubled the odds of remission in our cohort.

Long-term remission in patients with initial remission showed a delayed improvement of biochemical remission. Among secondary treatments in patients without initial remission, the lowest recurrence rate presented in patients undergoing repeat surgery, the necessity of which is highlighted in persistent cases with invasive tumors.^38,39^

### Limitations

Our study strengthens the insight into the patterns of care and results of treatment for acromegaly in China. The vast heterogeneity in the regional economy, insurance coverage, education, health literacy, and accessibility of medical care, however, may limit the generalizability of these results to all acromegaly patients in China. The self-reported nature of the CAPA database further contributed to a significant burden of missing data at extent of resection, detailed pathological results (Ki-67, somatostatin receptor subtype 2 expression, AIP expression, and granularity), dates of secondary treatment, and continuous follow-ups. Although we have the last follow-up record, which might underestimate the time to recurrence and therefore limit us to further investigate predictors of long-term outcomes. Finally, as evidenced by the low median age of our cohort, the majority of patients who contributed to this database through online submission were young, likely reflecting age-related biases in adopting an electronic reporting platform. Strategies to expand access and ease of data accrual from more diverse populations, both in demographics and age, may further diversify the value of this clinical database in the future.

## Conclusions

We present the largest nation-wide analysis of clinical features and surgical outcomes of acromegaly patients in China, driven from a patient-reported database. We highlight that higher preoperative GH levels, giant adenomas, and greater extent of tumor invasiveness are associated with a lower likelihood of biochemical remission at 3-months after surgery, while preoperative medical therapy increases the chance of remission. Importantly, we have established the effectiveness and potential of a patient-reported database in advancing neuroendocrinological research.

## Data Availability

To protect the privacy of participated patients, the anonymized data not presented within this article will only be available upon request from any qualified investigators.

## Acknowledgments

The authors would like to thank all the patients and families and their providers who collaborated with the establishment of CAPA database. We are grateful for the collaboration with the Chinese Association of Patients with Acromegaly (CAPA). We also thank Mr. Hao Luo (Warwick Business School, United Kingdom) for creating the China heat-map for the geographic distribution of acromegaly patients.

## Author Contributions

Conception and design: WL Chen, HJ Wang, and WL Bi. Acquisition of data: WL Chen and HJ Wang. Analysis and Interpretation of data: all authors. Drafting the article: S Yao, S Tavakol, and WL Bi. Critically revising the article: S Yao, S Tavakol, F Akter, MP Catalino, ER Laws, WL Bi, and HJ Wang. Reviewed submitted version of manuscript: all authors. Approved the final version of manuscript on behalf of all authors: HJ Wang. Statistical analysis: S Yao, S Tavakol, WL Bi. Graphic visualization: S Yao. Clinical diagnosis and radiological identification: WL Chen, J Luo, and ZG Mao. Administrative/technical/material support: WL Chen, ZG Mao, YH Zhu, and HJ Wang. Study supervision: HJ Wang and WL Bi.

## ABBREVIATIONS

ACTH: adrenocorticotropic hormone
CAPA: China Acromegaly Patient Association
CI: confidence interval
CT: computed tomography
DA: dopamine agonists
FDR: family discovery rate
GH: growth hormone
ICA: internal carotid artery
IGF-1: insulin-like growth factor 1
LH: luteinizing hormone
IQR: interquartile range
MRI: high-resolution magnetic resonance imaging
OR: odds ratio
PRL: prolactin (PRL)
FSH: follicle stimulating hormone
TSH: thyroid-stimulating hormone
VIF: variance influence factor
SD: standard deviation
SRLs: somatostatin receptor ligands

## Figure Legends

Supplemental Material S1. The geographic distribution of acromegaly patients in the China Acromegaly Patient Association (CAPA) database between 1998 and 2018, across 23 provinces, 4 autonomous regions, 4 municipalities, and 2 Special Administrative Regions (Hong Kong and Macau) in China.

